# COVID-19: An analysis of social media and research publication activity during the early stages of the pandemic

**DOI:** 10.1101/2020.12.20.20248517

**Authors:** Sonia L. Taneja, Monica Passi, Sumona Bhattacharya, Samuel A. Schueler, Sandeep Gurram, Christopher Koh

**Affiliations:** Digestive Diseases Branch, National Institutes of Diabetes and Digestive and Kidney Diseases, National Institutes of Health, Bethesda, Maryland; Urologic Oncology Branch, National Cancer Institute, National Institutes of Health, Bethesda, Maryland; Liver Diseases Branch, National Institutes of Diabetes and Digestive and Kidney Diseases, National Institutes of Health, Bethesda, Maryland

**Keywords:** coronavirus, COVID-19, social media, gastroenterology, SARS-CoV-2

## Abstract

**Objectives:** The COVID-19 pandemic has highlighted the importance of rapid dissemination of scientific and medical discovery. Social media (SoMe) has become an invaluable platform in science and medicine. This study analyzed activity of SoMe (Twitter), preprints, and publications related to COVID-19 and gastroenterology (GI) during the COVID-19 pandemic.

**Methods:** Data from Twitter, preprint servers and PubMed was collected and analyzed from December 2019 through May 2020. Global and regional geographic and gastrointestinal organ specific social media trends were compared to preprint and publication activity; any associations were identified.

**Results:** Over the 6-month period, there were 73,079 tweets from 44,609 users, 7,164 publications, and 4,702 preprints. Twitter activity peaked during March while preprints and publications peaked in April 2020. Strong correlations were identified between Twitter and both preprints and publications activity (p<0.001 for both). While COVID-19 data across the 3 platforms concentrated on pulmonology/critical care, the majority of GI tweets pertained to pancreatology, most publications focused on hepatology, and most preprints covered hepatology and luminal GI (LGI). There were significant associations between Twitter activity and research for all GI subfields (p=0.009 for LGI, p=0.006 for hepatology and IBD, p=0.007 for endoscopy), except pancreatology (p=0.2). Twitter activity was highest in the US (7,331 tweets) whereas PubMed activity was highest in China (1,768 publications).

**Conclusions:** The COVID-19 pandemic has highlighted the utility of SoMe as a vehicle for disseminating scientific information during a public health crisis. Scientists and clinicians should consider the use of SoMe in augmenting public awareness of their scholarly pursuits.

## INTRODUCTION

The current coronavirus disease 2019, abbreviated as COVID-19, is caused by the novel SARS-CoV-2 virus. It emerged into public view in December 2019 and has resulted in a pandemic that has affected six continents and continues to indiscriminately affect individuals of all ages, races and ethnicities. According to the World Health Organization, there has been more than 33,000,000 confirmed cases of COVID-19 globally, including more than 1,000,000 deaths reported by the end of September 2020 ^1^. While initial experiences related to COVID-19 primarily described respiratory complications, reports of gastrointestinal (GI) involvement became more evident with increased clinical experience ^2^.

Although the degree of GI involvement with COVID-19 was uncertain based on early published experiences, it was postulated that this could be substantial due to the identification of the SARS-CoV-2 entry mechanism utilizing the angiotensin 2 (ACE2) receptor pathway, which is found throughout the GI tract, liver, and pancreas. Given the pathogen’s similarities to SARS-CoV (SARS) and Middle East Respiratory Syndrome (MERS), investigators suspected that prior experiences with these preceding viruses could provide insight into the current pandemic. Thus, GI luminal manifestations, the involvement of the liver and pancreas, and the management of unique GI patient populations all became areas of clinical and research interest ^3-5^.

Considering the rapid spread of COVID-19 along with the interruption of healthcare services across multiple fronts, the importance of disseminating information in a timely and efficient manner became even more apparent. The publishing of international experience with COVID-19 along with frequent updates in clinical guidance documents have assisted the GI community in managing this novel disease ^6-9^. Furthermore, in an effort to mitigate the spread of infection, endoscopists have encountered significant changes to endoscopic practices by adopting new pre-procedure regulations, use of enhanced personal protective equipment, and the rearrangement of endoscopy units to facilitate social distancing ^10, 11^. Additionally, with the implementation of national “lockdowns,” the ability to share clinical experiences, analyze medical data and disseminate management strategies for COVID-19 has become reliant on electronic media ^12^. During these unprecedented times, the medical community has increasingly utilized social media (SoMe) (i.e. Twitter©, Facebook, TikTok) for communication and to facilitate interdisciplinary discussion ^13, 14^.

SoMe platforms, such as Twitter©, are microblogging and social networking services whereby scientific and medical communities can share information and achievements. Compared to other professions, the healthcare community has been relatively reluctant in utilizing SoMe for professional purposes related to concerns on its potential impact on employment, medicolegal liability, and relationships among patients and colleagues ^15-17^. Nonetheless, as these platforms continue to gain global acceptance and utilization, the ability to collect and analyze data from SoMe platforms have become essential in understanding healthcare related needs, shifting public health interests, and highlighting areas for further medical study ^18^. In this study, we aimed to explore COVID-19-related SoMe activity pertaining to the fields of Gastroenterology and Hepatology, over a six-month period by utilizing the Twitter© platform and to assess its impact on healthcare – specifically related to the dissemination of medical information and clinical practice. Furthermore, we aimed to compare SoMe to more traditional sources such as medical journal publications and preprint repositories, as vehicles in communicating medical information.

## MATERIALS AND METHODS

### Data collection for COVID-19-related Twitter activity

Data was collected utilizing the publicly available Twitter© analytics platform, Symplur Signals© (Symplur LLC, USA, www.symplur.com). Symplur Signals© is a healthcare social media (HCSM) analytics platform that utilizes algorithms with natural language processing to provide in-depth information on Twitter© activity. Data was collected from topics related to COVID-19 via the use of specific searches categorizing topics by organ system (see supplemental section). Data was captured over the first six months of the COVID-19 pandemic, from December 1, 2019 until May 31, 2020. In an effort to capture the longitudinal evolution of SoMe usage during this time period, each month was split into half (day 1 to day 15 and day 16 to end of the month). Data collected included: total number of tweets and retweets, total number of impressions, total number of users and user data including place of origin (by country globally and by state within the US). The ratios of tweets per Twitter© user and impressions per tweet were also calculated. Definitions of these terms can be found in the supplemental section.

### Data collection for COVID-19 related preprints and publications

Preprint articles are research manuscripts shared publicly before peer-review, which allows for the rapid dissemination of information, thereby helping to inform policy and clinical practice in a timely manner. Preprint repositories have gained considerable attention over the course of the pandemic and have been increasingly utilized for the dissemination of crucial pandemic science. Preprint articles related to COVID-19 were identified using two popular preprint servers for coronavirus biomedical research: MedRxiv (https://www.medrxiv.org/) and BioRxiv (https://www.biorxiv.org/). Specific search terms (see supplemental section) were utilized to identify and extract COVID-19 preprint articles for each half-month period over the six-month study period for comparison to Twitter data. A follow-up review of preprint articles pertaining to COVID-19 and GI that ultimately resulted in formal peer-reviewed journal publications was performed for the month of July 2020.

For the analysis of peer reviewed publications, the PubMed-National Center for Biotechnology Information (NCBI) database (https://pubmed.ncbi.nlm.nih.gov/) was utilized to identify all publications pertaining to COVID-19 over the six-month study period. The specific search terms used can be found in the supplemental section. All citations resulting from PubMed searches were recorded, and search results were further filtered by half-month time intervals, identical to the Twitter and pre-print search methods for the purposes of comparison. For both preprints and publications, articles were further subgrouped by organ system topic. Duplicate publications from separate searches were individually reviewed and recategorized into the most appropriate subject group, thereby eliminating the potential for publications to be accounted for more than once. Finally, for each publication, the geographical location of the first author’s institution was recorded.

### Analysis of social media, preprint, and publication activity

Overall trends in Twitter©, publications and preprint activity were analyzed and compared for temporal relationships with regards to peak activity, as well as by geographic locations demonstrating the most activity for each modality. Trends in activity over time were noted for each modality overall, as well as by specific organ-system topic (see supplemental section). Summary statistics of baseline data for tweets, impressions, preprints and PubMed publications are presented as frequencies for categorical data unless otherwise specified. Spearman’s rank-order correlation was performed to determine the relationships between Twitter activity (tweets and impressions) and PubMed publications overall, by organ system and geographical location as well as Twitter activity and preprint articles overall and by organ system. Analysis was performed using STATA 15 (StataCorp, LLC) (College Station, TX). Statistical significance was set at p<0.05. All authors had access to the study data and reviewed and approved the final manuscript.

## RESULTS

### COVID-19 related publications and Twitter© activity trends

Over the six-month study period from December 1, 2019 through May 31, 2020, 73,079 tweets were identified from a total of 44,609 users, generating 207,039,610 impressions on the topic of COVID-19. During this same period, 7,164 publications were identified to be indexed in PubMed along with 4,702 preprints archived in the MedRxiv and BioRxiv repositories pertaining to COVID-19. The overall summary of Twitter and publication activity by half-month time interval is shown in Table 1. Tweets on the topic of COVID-19 did not appear until the latter half of January 2020, which resulted in 245 original tweets. Twitter activity progressively increased thereafter and peaked during March 16-31, 2020 with 20,660 original tweets before gradually decreasing over the remaining study interval (Figure 1).

**Table 1.**
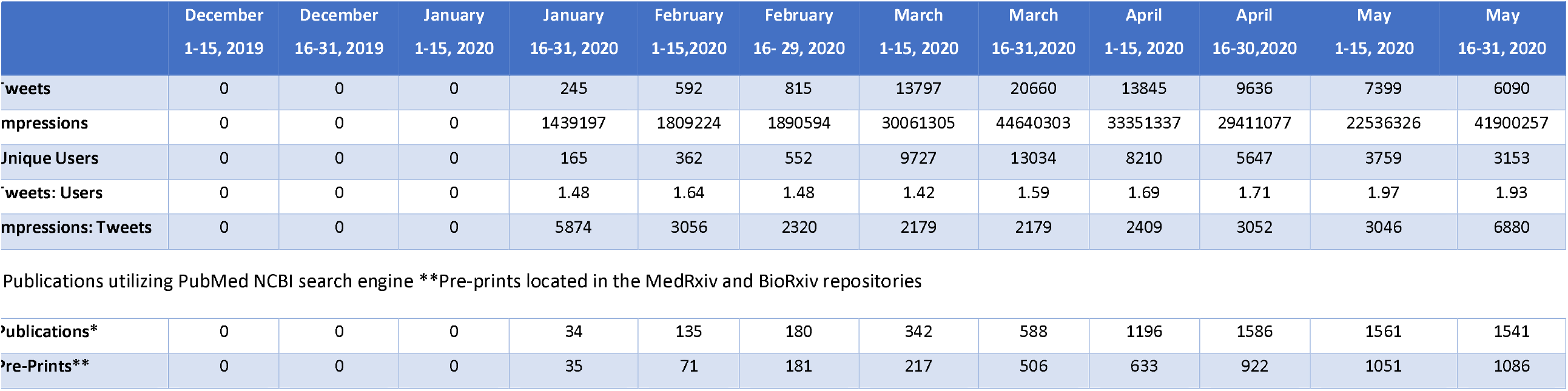
Summary of COVID-19 related Twitter, Publication and Pre-print Data From December 2019 through May 2020, Per Half-Month Intervals.

|  | December<br>1-15, 2019 | December<br>16-31, 2019 | January<br>1-15, 2020 | January<br>16-31, 2020 | February<br>1-15,2020 | February<br>16- 29, 2020 | March<br>1-15, 2020 | March<br>16-31,2020 | April<br>1-15, 2020 | April<br>16-30,2020 | May<br>1-15, 2020 | May<br>16-31, 2020 |
| --- | --- | --- | --- | --- | --- | --- | --- | --- | --- | --- | --- | --- |
| Tweets | 0 | 0 | 0 | 245 | 592 | 815 | 13797 | 20660 | 13845 | 9636 | 7399 | 6090 |
| Impressions | 0 | 0 | 0 | 1439197 | 1809224 | 1890594 | 30061305 | 44640303 | 33351337 | 29411077 | 22536326 | 41900257 |
| Unique Users | 0 | 0 | 0 | 165 | 362 | 552 | 9727 | 13034 | 8210 | 5647 | 3759 | 3153 |
| Tweets: Users | 0 | 0 | 0 | 1.48 | 1.64 | 1.48 | 1.42 | 1.59 | 1.69 | 1.71 | 1.97 | 1.93 |
| Impressions: Tweets | 0 | 0 | 0 | 5874 | 3056 | 2320 | 2179 | 2179 | 2409 | 3052 | 3046 | 6880 |
| Publications utilizing PubMed NCBI search engine \*\*Pre-prints located in the MedRxiv and BioRxiv repositories |  |  |  |  |  |  |  |  |  |  |  |  |
| Publications* | 0 | 0 | 0 | 34 | 135 | 180 | 342 | 588 | 1196 | 1586 | 1561 | 1541 |
| Pre-Prints** | 0 | 0 | 0 | 35 | 71 | 181 | 217 | 506 | 633 | 922 | 1051 | 1086 |

**Figure 1.**
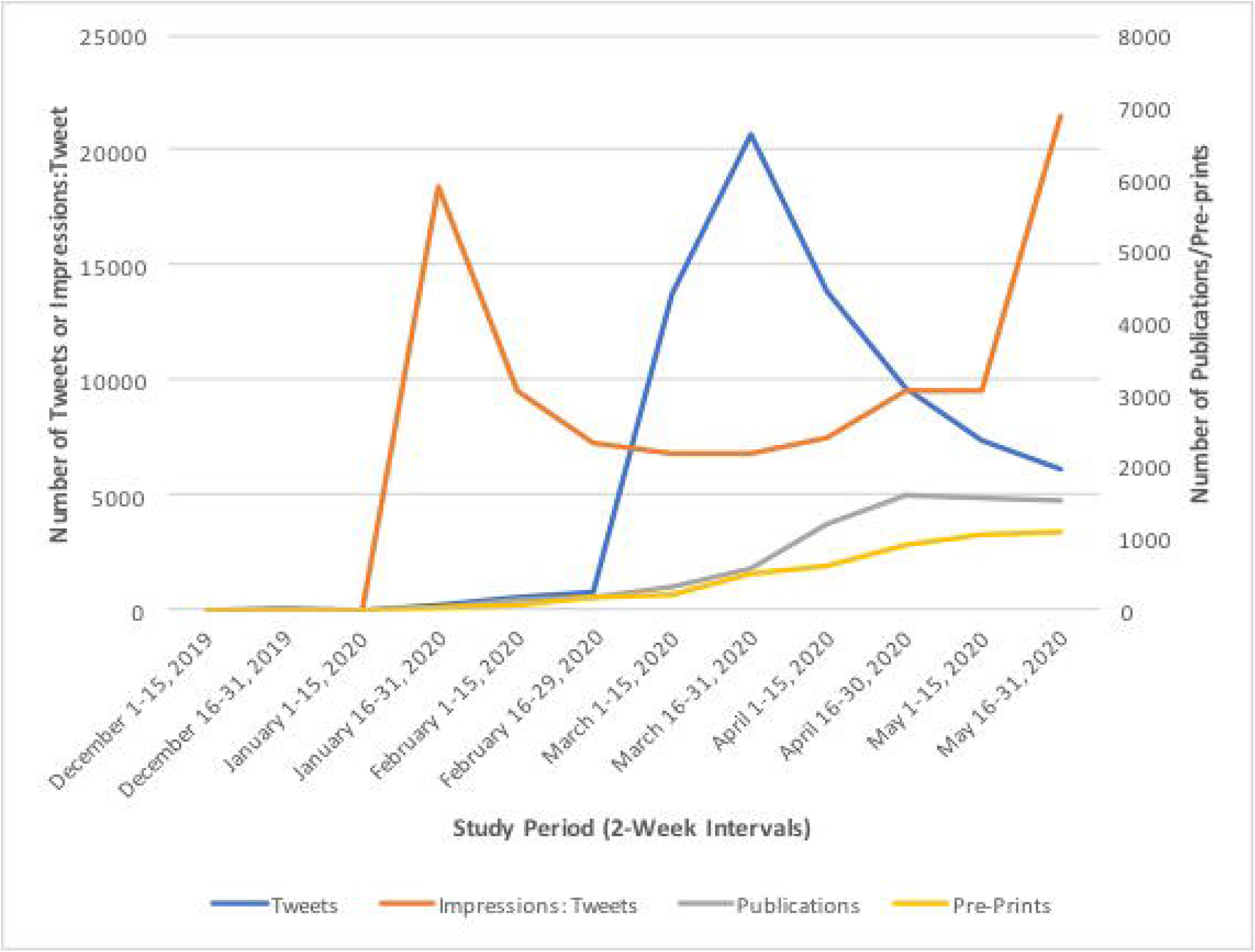
Trend of COVID 19 Tweets, Ratio of Impressions to Tweets, Publications and Pre-prints. Twitter activity from December 1, 2019 through May 2020 captured at half-month intervals utilizing Twitter© analytics platform, Symplur Signals ©. Pre-prints from MedRxiv and BioRxiv repositories also abstracted during this time period along with publications indexed in PubMed-NCBI database.

A similar pattern of activity was observed among the number of Twitter users which increased from 165 users to 13,034 users between January and the latter half of March 2020. Impressions followed a similar pattern with a peak during the second half of March 2020. Interestingly, a second peak in impressions was apparent during the latter half of May, which was not observed with number of tweets and Twitter users (Figure 2). On average, the number of tweets per Twitter user ranged from 1.48 to 1.97. Impressions generated per tweet, while initially high (5,874 impressions/tweet) in the latter half of January, did not peak until the latter half of May (6,880 impressions/tweet). Temporal trends are further detailed in Figure 1.

**Figure 2.**
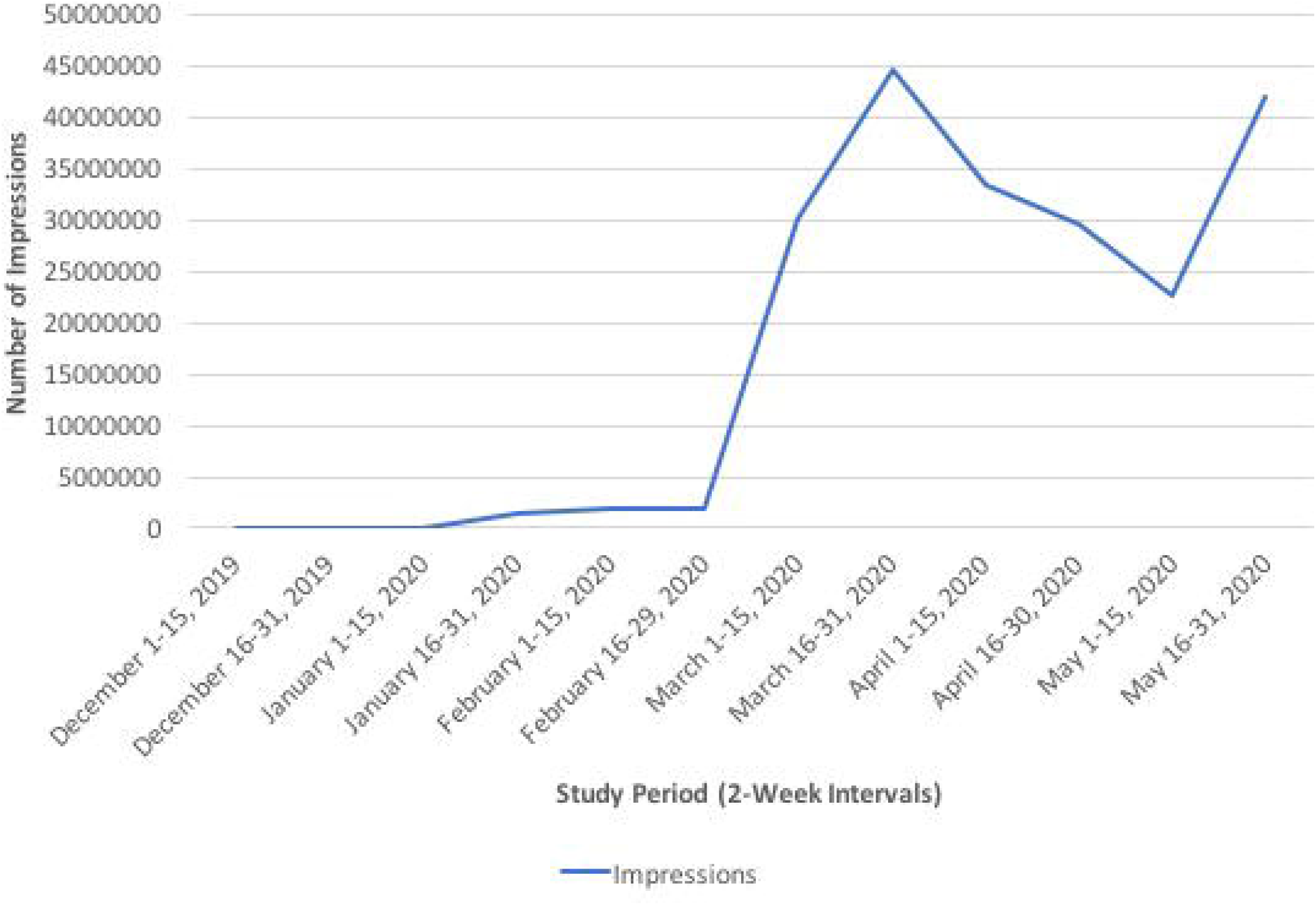
Trend of COVID-19 Twitter Impressions. Number of Impressions generated from Twitter © from December 1, 2019 through May 2020 captured at half-month intervals utilizing the Twitter© analytics platform, Symplur Signals ©.

Scientific COVID-19 related articles indexed in PubMed as well as preprints in MedRxiv and BioRxiv followed a similar trajectory as Twitter activity, with both the first peer-reviewed manuscripts and the first preprints appearing during the second half of January 2020 ^19^. However, unlike Twitter activity which peaked in the second half of March 2020, publications and preprints reached peak activity around the second half of April 2020. Notably, we observed a parallel rise in the number of preprints and PubMed publications (Table 1, Figure 1).

A moderately strong correlation was demonstrated between Twitter activity (i.e. number of tweets) and number of PubMed publications, as well as between Twitter activity and number of preprints over the entire study duration (r_s_: 0.58, p<0.001, for both; Table 2). Similarly, there was a moderately strong association between number of Twitter impressions and PubMed publications (r_s_: 0.56, p<0.001), as well as between the number of Twitter impression and preprints (r_s_: 0.54, p<0.001) (Table 3).

**Table 2.** Correlation Between Twitter Activity and PubMed Publications and between Twitter Activity and Pre-Prints by Organ System.

| Organ System | Tweets & PubMed Publications<br>($\rho^*$) | P-value | Tweets & Pre-Prints ($\rho^*$) | P-value |
| --- | --- | --- | --- | --- |
| **Overall Trend** | 0.58 | *<0.001* | 0.57 | *<0.001* |
| **Trend by Organ System** |  |  |  |  |
| *Pulmonology/Critical Care* | 0.8 | *0.002* | 0.8 | *0.003* |
| *LGI* | 0.7 | *0.009* | 0.6 | *0.03* |
| *Hepatology* | 0.7 | *0.006* | 0.7 | *0.009* |
| *IBD* | 0.7 | *0.006* | 0.5 | 0.07 |
| *Pancreatology* | 0.4 | 0.2 | 0.4 | 0.3 |
| *GI Endoscopy* | 0.7 | *0.007* | 0.7 | *0.02* |

\* $\rho$ : correlation coefficient

**Table 3.**
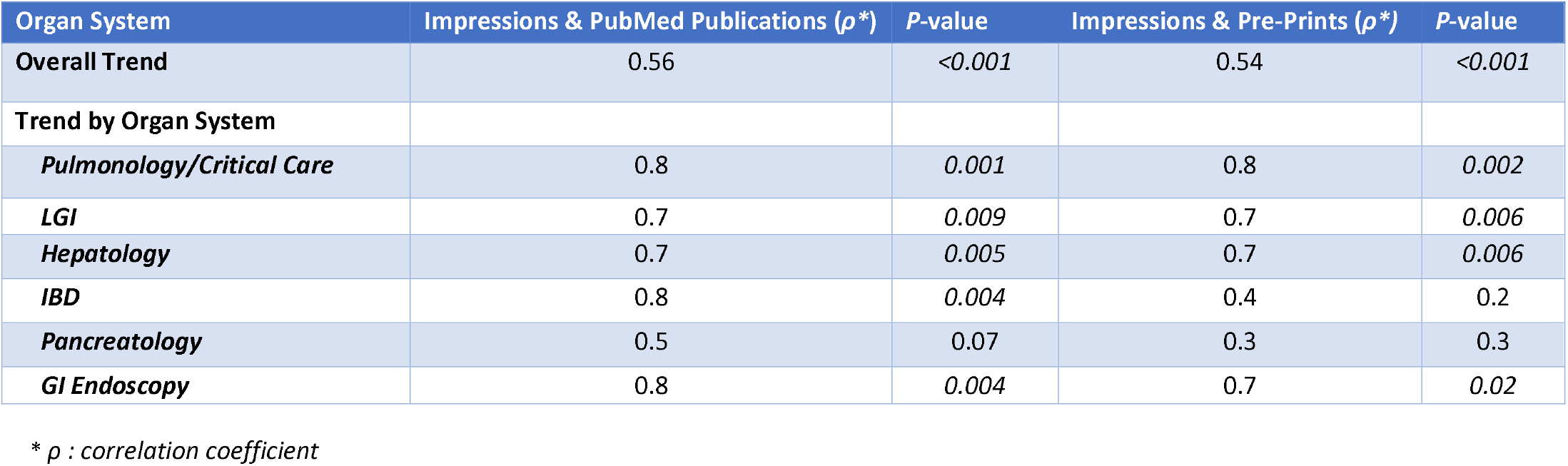
Correlation Between Twitter Impressions and PubMed Publications & Between Twitter Impressions and Pre-Prints by Organ System.

### COVID-19 Twitter, publication, and preprint content by organ system topic

Analyzing Twitter, publication, and preprint data pertaining to the effects of COVID-19 on specific organ system topics are outlined in Table 4. The majority of COVID-19 related tweets, publications and pre-prints covered the topic of pulmonology/critical care (80.4%, 93.8% and 99.0%, respectively). Within the field of GI, the majority of tweets were on the topics of pancreatology (7.9% of all tweets), followed by luminal GI (LGI) (4.5%), inflammatory bowel diseases (IBD) (3.9%), gastrointestinal (GI) endoscopy (2.4%) and hepatology (0.8%). Alternatively, regarding GI-related publications, the majority of articles were on the topic of hepatology (3.3%) followed by GI endoscopy (1.5%), LGI (0.9%), IBD (0.4%), and pancreatology (0.1%). Preprint publications were primarily on the topics of LGI and hepatology (0.4% each) and IBD (0.1%) (Table 4).

**Table 4.** Summary of COVID 19 related Tweets, Publications and Pre-print Activity by Organ System per Half-Month Intervals.

|  | Pulmonology/<br>Critical Care | Luminal Gastroenterology<br>(LGI) | Hepatology | IBD | Pancreatology | GI Endoscopy |
| --- | --- | --- | --- | --- | --- | --- |
| **Dec. 1 – Dec. 15, 2019** |  |  |  |  |  |  |
| **Tweets** | 0 | 0 | 0 | 0 | 0 | 0 |
| **Publications** | 0 | 0 | 0 | 0 | 0 | 0 |
| **Pre-Prints** | 0 | 0 | 0 | 0 | 0 | 0 |
| **Dec. 16 – Dec. 31, 2019** |  |  |  |  |  |  |
| **Tweets** | 0 | 0 | 0 | 0 | 0 | 0 |
| **Publications** | 0 | 0 | 0 | 0 | 0 | 0 |
| **Pre-Prints** | 0 | 0 | 0 | 0 | 0 | 0 |
| **Jan. 1- Jan. 15, 2020** |  |  |  |  |  |  |
| **Tweets** | 0 | 0 | 0 | 0 | 0 | 0 |
| **Publications** | 0 | 0 | 0 | 0 | 0 | 0 |
| **Pre-Prints** | 0 | 0 | 0 | 0 | 0 | 0 |
| **Jan.15- Jan 31, 2020** |  |  |  |  |  |  |
| **Tweets** | 228 | 4 | 0 | 2 | 11 | 0 |
| **Publications** | 34 | 0 | 1 | 0 | 0 | 0 |
| **Pre-prints** | 0 | 0 | 0 | 0 | 0 | 0 |
| **Feb. 1- Feb. 15, 2020** |  |  |  |  |  |  |
| **Tweets** | 556 | 6 | 0 | 14 | 11 | 5 |
| **Publications** | 134 | 0 | 1 | 0 | 0 | 0 |
| **Pre-prints** | 70 | 0 | 1 | 0 | 0 | 0 |
| **Feb. 16 – Feb. 29, 2020** |  |  |  |  |  |  |
| **Tweets** | 740 | 2 | 0 | 2 | 50 | 21 |
| **Publications** | 174 | 1 | 5 | 0 | 0 | 0 |
| **Pre-prints** | 177 | 3 | 1 | 0 | 0 | 0 |
| **Mar. 1 – Mar. 15, 2020** |  |  |  |  |  |  |
| **Tweets** | 10406 | 206 | 201 | 1500 | 1200 | 284 |
| **Publications** | 331 | 1 | 8 | 1 | 0 | 1 |
| **Pre-prints** | 214 | 1 | 1 | 0 | 1 | 0 |
| **Mar. 16 – Mar. 31, 2020** |  |  |  |  |  |  |
| **Tweets** | 16489 | 1769 | 165 | 502 | 1200 | 535 |
| **Publications** | 567 | 2 | 12 | 3 | 0 | 4 |
| **Pre-prints** | 497 | 3 | 4 | 1 | 0 | 1 |
| **Apr. 1 – Apr. 15, 2020** |  |  |  |  |  |  |
| **Tweets** | 11674 | 399 | 18 | 206 | 1300 | 248 |
| **Publications** | 1131 | 11 | 31 | 1 | 1 | 21 |
| **Pre-prints** | 629 | 2 | 2 | 0 | 0 | 0 |
| **Apr. 16 – Apr. 30, 2020** |  |  |  |  |  |  |
| **Tweets** | 7728 | 327 | 10 | 216 | 924 | 431 |
| **Publication** | 1500 | 12 | 44 | 4 | 2 | 24 |
| **Pre-prints** | 908 | 8 | 4 | 1 | 0 | 1 |
| **May 1 – May 15, 2020** |  |  |  |  |  |  |
| **Tweets** | 5945 | 403 | 143 | 205 | 559 | 144 |
| **Publications** | 1439 | 19 | 58 | 11 | 3 | 31 |
| **Pre-prints** | 1046 | 1 | 3 | 1 | 0 | 0 |
| **May 16 – May 31, 2020** |  |  |  |  |  |  |
| **Tweets** | 5026 | 202 | 46 | 171 | 549 | 96 |
| **Publications** | 1405 | 17 | 75 | 10 | 4 | 30 |
| **Pre-prints** | 1082 | 2 | 2 | 0 | 0 | 0 |

\*Publications utilizing PubMed NCBI search engine \*\*Pre-prints located in the MedRxiv and BioRxiv repositories

### Pulmonology/Critical Care

Approximately 59,000 tweets were on the topic of pulmonology/critical care and COVID-19 over the entire study period. The most significant increase in tweets on this topic occurred between February and March 2020, with a nearly 22-fold increase, and an ultimate peak in activity observed in the latter half of March (16,489 tweets) (Table 4).

A total of 6,713 peer-reviewed articles on the topic of pulmonology/critical care and COVID-19 were indexed in PubMed during the six-month study period, and first appeared in January. The most significant increase in number of publications on COVID-19 and pulmonology/critical care was observed between the latter half of March and the start of April 2020, with a two-fold increase. As compared to peer-reviewed publications, there were approximately one-third fewer preprint articles (4,567) in MedRxiv and BioRxiv related to COVID-19 and pulmonology/critical care identified during the study period. On the topic of pulmonology/critical care and COVID-19, the longitudinal trend in pre-print article availability appears to have paralleled publications indexed in PubMed; however, for pre-print articles, the most significant rise was seen two weeks prior to that observed with PubMed publications, specifically between the first and second half of March (Table 4).

There was a strong correlation between both the number of tweets and peer-reviewed publications as well as between the number of tweets and preprints on the topic of COVID-19 and pulmonology/critical care (r_s_: 0.8, p=0.002 and r_s_: 0.8, p=0.003, respectively) (Table 2). Similarly, there was a strong correlation between both pulmonology/critical care-related Twitter impressions and publications (r_s_:0.8, p=0.001) as well as between Twitter impressions and preprints (r_s_: 0.8, p=0.002) (Table 3).

### Gastroenterology Topics

A total of 14,285 tweets concerning the field of GI and COVID-19 (encompassing subspecialty fields of LGI, IBD, hepatology, GI endoscopy and pancreatology) were identified during the entire study period (**Table 4**). Among all tweets recorded during the six-month study period, 19.6% were on the topic of COVID-19 and GI. The longitudinal trend in number of GI related tweets (including subspecialty GI fields) paralleled that observed with pulmonology/critical care related tweets, with an approximate 45-fold increase in number of tweets spanning the latter half of February and peaking in the second half of March. When further stratified by subspecialty field, the majority of COVID-19 GI related tweets were on the topic of pancreatology (40.6%), followed by LGI (23.2%), IBD (19.7%), GI endoscopy (12.3%) and hepatology (4.1%).

A total of 449 peer-reviewed publications related to COVID-19 and GI were identified in PubMed during the entire study period. In contrast to Twitter activity, the majority of these publications were on the topic of hepatology (52.3%) followed by GI endoscopy (24.6%), LGI (14.2%), IBD (6.7%) and pancreatology (2.2%). Similar to Twitter activity, PubMed publications on the topics of LGI and hepatology first appeared in the latter half of January 2020. The most significant increase in COVID-19 liver related publications was observed between the latter half of March into early April, with an over 2.5-fold increase in the number of publications on this topic (12 versus 31 articles). LGI publications, which first appeared in the latter half of February, significantly increased between the second half of March into early April, with a five-fold increase as detailed in Table 4.

There was a total of 45 COVID-19 and GI related preprints archived in MedRxiv and BioRxiv over the study period. On longitudinal analysis, the number of pre-prints on the topic of GI peaked in the latter half of April. When further stratified by subspecialty, unlike that observed with peer-reviewed publications, the majority of preprints covered LGI (46.7%), followed by hepatology (40%), IBD (6.7%), GI endoscopy (4.4%) and pancreatology (2.2%) (Table 4).

Similar to pulmonology/critical care related content, there was a strong correlation between tweets and peer-reviewed publications (r_s_: 0.6, p=0.03) as well as between tweets and preprints on the topic of LGI (r_s_: 0.7, p=0.009). Additionally, a strong correlation was identified between both the number of tweets and peer-reviewed publications (r_s_: 0.7, p=0.006) as well as between tweets and preprints on the topic of COVID-19 and hepatology (r_s_:0.7, p=0.009). A similarly strong correlation was demonstrated between the number of tweets and PubMed publications on the topic of GI endoscopy (r_s_: 0.7, p=0.007), the number of tweets and preprints regarding GI endoscopy (r_s_:0.7, p=0.02), and the number of tweets and peer-reviewed publications on the topic of COVID-19 and IBD (r_s_: 0.7, p=0.008). On the contrary, no associations were identified between tweets and peer-reviewed publications and preprints on the topic of COVID-19 and pancreatology (Table 2).

Regarding COVID-19 and LGI content, a strong correlation was identified between both Twitter impressions and peer-reviewed publications (r_s_:0.7, p=0.009) as well as between Twitter impressions and preprints (r_s_:0.7, p=0.006). Similarly, strong correlations were identified between Twitter impressions and peer-reviewed publications on the topics of COVID-19 and hepatology (r_s_:0.7, p=0.005), IBD (r_s_:0.8, p=0.004) and GI Endoscopy (r_s_:0.8, p=0.004). There was no correlation between Twitter impressions and publications on the topic of COVID-19 and pancreatology. In evaluating the association between Twitter impressions and preprints, there were also strong associations on the topics of hepatology (r_s_:0.7, p=0.006) and GI endoscopy (r_s_: 0.7, p=0.02), whereas there were no associations were found pertaining to the topics of IBD and pancreatology (Table 3).

### COVID-19 Twitter and publication content by geographical location

The top 5 countries with the most number of tweets over the six-month study period pertaining to COVID-19 included the US (33.0% of total tweets globally), followed by the United Kingdom (UK) (19.3%), Spain (6.8%), Canada (5.3%), and Australia (3.0%). Alternatively, China was the country generating the highest number of peer-reviewed publications indexed in PubMed (1,768) over the entire study period, followed by Italy (915), US (389), France (348), and India (303). Figure 3 illustrates the countries with the most Twitter (Fig. 3A) and peer-reviewed publication (Fig. 3B) activity. The top 20 countries with the highest number of tweets and the highest number of peer-reviewed publications are detailed in supplemental tables 2 and 3, respectively. There was a strong correlation between the number of Twitter and peer reviewed publications amongst both the US (r_s_: 0.8, p=0.005) and the UK (r_s_: 0.8, p=0.01) (Supplemental Table 1).

**Figure 3 A-D.**
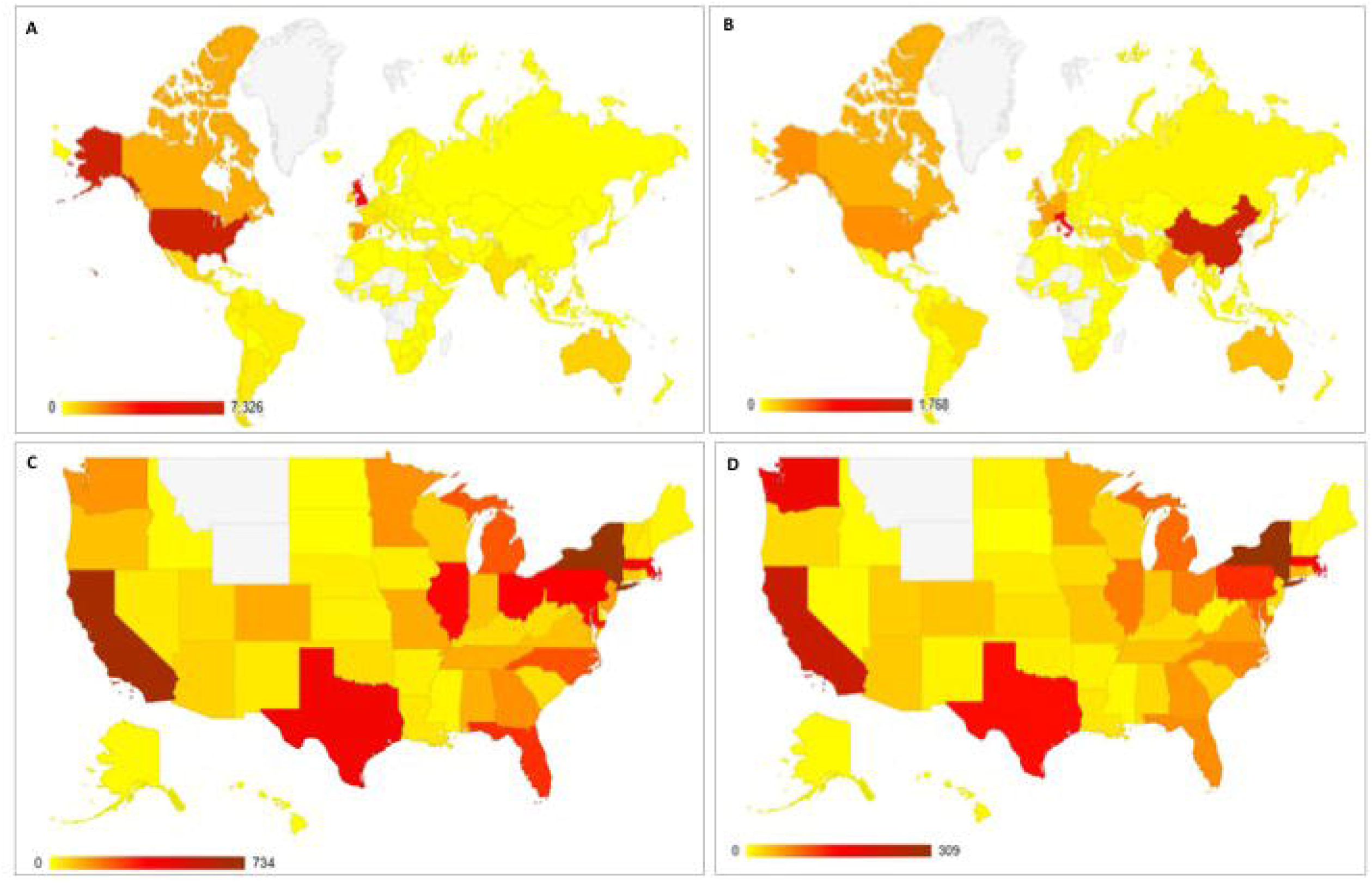
Global and United States (US) Heat Maps Illustrating Twitter and PubMed Publication Activity. Total number of COVID 19 related Tweets and publications as indexed in PubMed NCBI database over the 6 month study period (December 2019 through May 2020) represented globally (A (PubMed publications), B (Tweets)), as well as within the US (C (PubMed publications), D (Tweets)). Number represented on spectrum from least (yellow) to most amount (maroon), as detailed in the legend accompanying each map.

Within the US, when analyzing both Twitter activity and peer reviewed publications, New York state had the highest COVID-19 related activity (11% of tweets and 39.1% of publications) followed closely by California (10.3% of tweets and 36.6% of publications) during the entire study period. Figure 3 illustrates the US states with the most Twitter (Fig. 3C) and publication (Fig. 3D) activity. The top 20 US states with the highest number of Twitter and peer reviewed publications is detailed in Supplemental Tables 4 and 5, respectively.

### COVID-19 twitter content by user stakeholder designation/category

Twitter user data entered as the Twitter user’s self-designated healthcare stakeholder role was analyzed. For the topics of pulmonology/critical care, LGI, hepatology, IBD and GI endoscopy, the top two most active stakeholder categories were doctors/physicians and researchers/academic users. Alternatively, advocacy organizations and patient advocates were the most active stakeholder users for the topic of pancreatology. The top 15 most active users and their stakeholder roles categorized by Twitter activity for each organ system topic are further detailed in Supplemental Table 6.

## DISCUSSION

Since its emergence on the world stage in December 2019, the novel SARS-CoV-2 coronavirus has triggered an unparalleled global response in the fields of science, medicine, public health and technology. Considering this virus’ highly contagious nature, along with the paucity of knowledge and current absence of effective treatment modalities to combat the infection, the need for rapid sharing and dissemination of information has been paramount. In this study, we assessed the dissemination of COVID-19 related information via preprint services, formal peer reviewed publications, and through the global reach of social media via Twitter©.

Specifically, we observed that during the second half of March 2020, when COVID-19 was continuing to spread rapidly prompting various nations to go into lockdown, SoMe activity via Twitter was at its peak, with almost 7,000 impressions per tweet. Furthermore, we observed that Twitter activity strongly correlated with the availability of published, scientific data to the general public. Although COVID-19 has been predominantly linked with severe pulmonary complications, approximately 20% of the conversation on SoMe was related to the field of GI with specific discussion related to hepatology, GI endoscopy, LGI, IBD, and pancreatology. SoMe traffic was strongly associated with the availability of published data pertaining to all GI topics with the exception of pancreatology. Finally, in our analysis of data by geographic region of publication, activity was most prominent in regions most affected by the pandemic both globally (in regions where SoMe via Twitter is not banned) and within the U.S., with strong associations between SoMe and publication data.

In the longitudinal assessment of publication activity, we observed that peak SoMe activity pre-dated peak PubMed publications by approximately 30 days. One possible explanation for this lag interval is the technical delay that typically occurs after acceptance of a peer reviewed manuscript by a journal and prior to indexing in PubMed. It is worth highlighting that around the peak of PubMed publication activity, we also witnessed a parallel rise in the pre-print repository activity. Historically, the use of pre-print repositories has allowed researchers to “claim the space” or even to “publish first” in contentious areas of science and research. However, in the face of an evolving pandemic, preprint repositories are serving as a new mode of scientific communication, allowing for faster dissemination and communication of research and clinical findings related to COVID-19. In fact, in March 2020 when the WHO officially declared COVID-19 a pandemic, 8,830 biomedical preprints were published, a 142% increase from last year and MedRxiv page views have risen to 15 million a month, as compared to 1 million a month prior to the start of the pandemic ^20^. As of July 1, 2020, nearly half of all COVID-19 and GI related preprint articles have subsequently made their way into reputable scientific journals, further supporting this theory.

While pancreatic manifestations (e.g., acute pancreatitis) of COVID-19 have been less commonly reported in the literature as compared to other GI symptoms, we unexpectedly observed that Twitter activity on the topic of pancreatology was second to that of pulmonology/critical care. However, there was no statistically significant association between Twitter and publication and Twitter and pre-print activity on the topic of pancreatology, possibly attributable to the stakeholder data for pancreatology related tweets. Whereas stakeholder demographics fell largely under physicians, non-medical doctors, or researchers/academic users for each of the other GI subfields, for the topic of “Pancreas,” the most active stakeholders appeared to be advocacy organizations or patient advocates. Upon further review of the top 50 associated hashtags used under the “Pancreas” topic, those related to cystic fibrosis support and awareness groups were the most common. Patient advocacy organizations not only play an important part in espousing awareness of specific diseases and patient populations, but also serve an essential role in the dissemination of information on their behalf, typically in the form or raising awareness online, lobbying directly for change within government of other institutions, and via marketing and outreach. As advocacy groups more frequently use these alternative means for sharing information, this increase in Twitter activity may not translate to a proportionate rise in publication activity on the topic of COVID-19 and pancreatology as demonstrated in this study.

Notably, we observed that physicians, non-medical doctors, and scientific researchers constituted the lead stakeholder activity for SoMe use overall during the COVID-19 pandemic. This observation suggests that even during the early stages of the COVID-19 pandemic, SoMe became an increasingly sought-after tool for the purposes of communicating medical and scientific information. As the COVID-19 pandemic has disrupted the way research is ordinarily conducted, the scientific community has leveraged social media platforms to engage each other and as a means to collaborate on interdisciplinary research endeavors. Previous studies, performed during non-pandemic times, have demonstrated conflicting evidence as to the impact of SoMe on scientific publications related to citation impact, metrics and viewability ^21-24^. Our study has provided a unique opportunity to understand how SoMe can interact and facilitate research efforts during a pandemic in which clinical and scientific knowledge is rapidly evolving. Additionally, this study has provided an opportunity to increase our understanding of the influence of SoMe on the visibility of scientific research and scholarly productivity.

There are several limitations to our study worth noting. The first major limitation is that social media, such as Twitter, is not available in several countries, including China. This could help to explain China’s lead in publication activity as compared with other regions, as this is likely one of the primary modalities used in this country for disseminating information. Other social media platforms, including WeChat and Sina Weibo, are used in China, however information regarding the use of these other social media platforms within China is limited to date. Future studies are needed to assess SoMe activity on these alternative platforms and their association with publications as well as how they compared to the use of Twitter in other nations, such as the US, where Twitter is not banned. Secondly, although we were able to account for and re-assign duplicate publications for the various categorizations performed, the same was not able to be guaranteed for Twitter data. The very nature of tweets allows for other users to publish an original tweet to their account generating additional impressions (also known as a “retweet”). Therefore, limiting duplicate tweets may artificially decrease activity. More importantly, certain tweets, or even retweets, may have been assigned to more than one topic area by the Symplur software. Due to limitations with Symplur, we were unable to limit tweets to a single topic, and as such, this may have artificially boosted the overall number in certain organ system topics, thereby potentially skewing the results of our study.

In conclusion, this study demonstrates the productivity of social media (specifically, Twitter), pre-print, and publication methods for the rapid dissemination of information during an infectious global pandemic. As the world faces this unprecedented public health emergency, our study has reflected on shifting, worldwide trends from solely traditional methods of disseminating information (i.e., via publications) to more contemporary methods specifically among the GI community. SoMe tools, like Twitter, can be an effective method for educating and informing audiences in real time and via an interactive approach, a feat that cannot always be achieved with more conventional methods (i.e., scientific publications). The strong association that our study has demonstrated between Twitter activity, preprints and publications regarding COVID-19 and most GI subcategories further underscores the influence that social media may have on scholarly pursuits. The New Media Age has resulted in a number of novel avenues for the distribution of information, including Twitter©. It may be time for the medical and scientific communities to cultivate formal SoMe platforms as effective tools for data sharing and collaboration to augment existing modalities of archival publication.

## Supporting information

Supplemental Section

## Data Availability

All data referred to in the manuscript and note links is publicly available.

https://www.symplur.com

https://www.biorxiv.org/

https://pubmed.ncbi.nlm.nih.gov/

https://www.twitter.com

## Abbreviations

(COVID-19): coronavirus disease 2019
(GI): gastroenterology
(HCSM): healthcare social media
(IBD): inflammatory bowel disease
(LGI): luminal gastroenterology
(SARS-CoV-2): severe acute respiratory syndrome coronavirus 2
(SoMe): social media
(US): United States
(UK): United Kingdom

