## Supplemental Section for "COVID-19: An analysis of social media and research publication activity during the early stages of the pandemic"

**Search Terms used for Twitter, PubMed, and Pre-print Repositories**

Over the 6 month study period, data was recorded by half-month time intervals. The dates utilized for each search were: December 1 – 15, 2019; December 16 – 31, 2019; January 1 – 15, 2020; January 16 – 31, 2020; February 1 – 15, 2020; February 16 – 29, 2020; March 1 – 15, 2020; March 16 – 31, 2020; April 1 – 15, 2020; April 16 – 30, 2020; May 1 – 15, 2020; and May 16 – 31, 2020.

The following Twitter topics were searched: Pulmonology, Critical Care, Gastroenterology, Hepatology, Pancreas, Endoscopy, and Inflammatory Bowel Disease. Tweets within a given topic were filtered further to capture SARS-CoV-2 related tweets using the following keywords: coronavirus, covid19, covid-19, sarscov2, and SARS-CoV-2.

The following search terms were used to gather pre-print repository data: SARS-CoV-2, COVID-19, or Coronavirus. An additional search term was then applied to identify those preprints pertaining to the GI tract, including: gastroenterology, gastrointestinal, hepatology, liver, endoscopy, pancreas, and inflammatory bowel disease. Those articles not pertaining to the GI tract were classified under pulmonology/critical care given the respiratory predominant infectious nature of COVID-19.

The following search terms were utilized to perform PubMed literature review: coronavirus, COVID 19, COVID-19, Sarscov2, or Sars-cov-2. A second search term was then used to narrow the search to a specific organ system of interest including: pulmonary, pulmonology, lung, respiratory, critical care, ICU, gastroenterology, GI, endoscopy, liver, hepatology, inflammatory bowel disease or pancreas.

**Twitter Definitions**

The number of Tweets includes unique Tweets created as well as those that were shared by other users, also known as “retweets”. The number of users represents the number of unique Twitter accounts that created Tweets. The number of impressions is the number of users that a Tweet could have theoretically reached (based on the number of followers of the account that tweeted). Impressions generated are calculated by multiplying a Tweet by the number of Twitter users that follow the particular user account who produced the Tweet. The ratio of tweets per Twitter user was calculated by dividing the number of tweets for a particular half-month time interval by the number of users found during that same half-month period. Similarly, the ratio of impressions per tweet was calculated by dividing the number of impressions in a half-month time interval by the number of tweets found during that same half-month period. Finally, data pertaining to each Twitter account was also collected, specifically, location of the user and account type, both of which are self-reported. Account type refers to the self-designated healthcare stakeholder role that represented the Twitter user’s account (e.g., physician, professional organization, government organization, pharmaceutical company).
