## Supplemental Tables for "COVID-19: An analysis of social media and research publication activity during the early stages of the pandemic"

| Organ System | Twitter Activity & PubMed Publications (*ρ*)* | *P*-value |
| --- | --- | --- |
| Overall Trend | 0.50 | *<0.0001* |
| Trend by Country |  |  |
| *United States* | 0.8 | *0.005* |
| *United Kingdom* | 0.8 | *0.01* |
| *Australia* | 0.6 | 0.1 |
| *Spain* | 0.5 | 0.2 |
| *Italy* | 0.5 | 0.2 |
| *France* | 0.6 | 0.07 |

**Supplemental Table 1. Correlation Between Twitter Activity and PubMed Publications by Country**

*ρ****:*** *represents correlation coefficient.*

**Supplemental Table 2. Top 20 countries with the highest number of Twitter Activity Over 6-Month Study Period**

| Country | No. of Tweets |
| --- | --- |
| United States | 7331 |
| United Kingdom | 4229 |
| Spain | 1527 |
| Canada | 1174 |
| Australia | 673 |
| Malaysia | 643 |
| Mexico | 597 |
| Ireland | 470 |
| India | 460 |
| France | 357 |
| Saudi Arabia | 317 |
| Italy | 290 |
| Argentina | 262 |
| Colombia | 253 |
| Brazil | 249 |
| Chile | 238 |
| Germany | 227 |
| Ecuador | 226 |
| South Africa | 143 |
| Turkey | 130 |

**Supplemental Table 3. Top 20 countries with the highest number of Publications Indexed In PubMed NCBI Over 6-Month Study Period**

| Country | No. of Publications |
| --- | --- |
| China | 1768 |
| Italy | 915 |
| United States | 389 |
| France | 348 |
| India | 303 |
| Germany | 287 |
| Canada | 269 |
| United Kingdom | 257 |
| Australia | 231 |
| Hong Kong | 191 |
| Spain | 184 |
| Switzerland | 177 |
| Iran | 153 |
| Singapore | 150 |
| Netherlands | 149 |
| Japan | 142 |
| Korea | 128 |
| Brazil | 112 |
| Saudi Arabia | 105 |
| Israel | 94 |

**Supplemental Table 4. Top 20 US States with the Highest Number of Twitter Activity Over the 6-Month Study Period**

| US State | No. of Tweets |
| --- | --- |
| New York | 734 |
| California | 688 |
| Texas | 419 |
| Massachusetts | 377 |
| Pennsylvania | 376 |
| Ohio | 370 |
| Illinois | 357 |
| Maryland | 345 |
| Florida | 300 |
| North Carolina | 247 |
| Michigan | 242 |
| Georgia | 160 |
| Minnesota | 157 |
| Washington | 153 |
| New Jersey | 135 |
| Colorado | 123 |
| Missouri | 120 |
| Tennessee | 116 |
| Alabama | 113 |
| Virginia | 91 |

**Supplemental Table 5. Top 20 US States with the Highest Number of Publications Over the 6-Month Study Period**

| US State | No. of Publications |
| --- | --- |
| New York | 309 |
| California | 236 |
| Washington State | 182 |
| Massachusetts | 160 |
| Pennsylvania | 127 |
| Maryland | 92 |
| Michigan | 89 |
| Illinois | 78 |
| Ohio | 77 |
| North Carolina | 72 |
| Florida | 69 |
| Georgia | 61 |
| Virginia | 56 |
| Minnesota | 53 |
| Connecticut | 42 |
| Tennessee | 37 |
| Colorado | 36 |
| Missouri * | 35 |
| Indiana * | 35 |
| Arizona | 33 |

*equal number of publications

**Supplemental Table 6. The 15 Most Active Twitter Users’ Self-Designated Healthcare Stakeholder Roles Categorized by Number of Tweets Per Organ System Topic**

| **Stakeholder** | **# of tweets for Pancreatology** | **Stakeholder** | **# of tweets for Pulmonology/Critical Care** | **Stakeholder** | **# of tweets for LGI** |
| --- | --- | --- | --- | --- | --- |
| Org. Advocacy | 113 | Doctor | 4611 | Doctor | 387 |
| Org. Advocacy | 87 | Doctor | 3407 | Researcher/Academic | 116 |
| Org. Advocacy | 73 | Doctor | 3076 | Org. Advocacy | 115 |
| Individual Non-Health | 68 | Doctor | 2031 | Doctor | 108 |
| Org. Advocacy | 64 | Doctor | 1829 | Org. Advocacy | 106 |
| Org. Advocacy | 60 | Doctor | 1303 | Professional Org. | 91 |
| Patient Advocate | 60 | Researcher/Academic | 1137 | Doctor | 89 |
| Doctor | 48 | HCP | 957 | Doctor | 87 |
| Individual Non-Health | 48 | Org. Advocacy | 915 | Doctor | 67 |
| Patient Advocate | 45 | Researcher/Academic | 837 | Org. Advocacy | 66 |
| Patient Advocate | 43 | HCP | 745 | Physician | 65 |
| Doctor | 38 | Org. Advocacy | 671 | Individual Non-Health | 63 |
| Org. Advocacy | 32 | Researcher/Academic | 656 | Org. Advocacy | 57 |
| Individual Other Health | 30 | Org. Advocacy | 625 | Professional Org. | 54 |
| Individual Non-Health | 30 | Researcher/Academic | 589 | Org. Other Healthcare | 46 |

| **Stakeholder** | **# of tweets for hepatology** | **Stakeholder** | **# of tweets for IBD** | **Stakeholder** | **# of tweets for GI endoscopy** |
| --- | --- | --- | --- | --- | --- |
| Doctor | 75 | Doctor | 361 | Doctor | 203 |
| Doctor | 74 | Researcher/Academic | 145 | Doctor | 141 |
| Researcher/Academic | 31 | Patient Advocate | 133 | Doctor | 129 |
| Doctor | 26 | Doctor | 110 | Doctor | 89 |
| Researcher/Academic | 23 | Physician | 106 | Researcher/Academic | 55 |
| Org. Advocacy | 19 | Org. Advocacy | 96 | Org. Advocacy | 51 |
| Org. Advocacy | 18 | Doctor | 80 | Org. Advocacy | 41 |
| Physician | 15 | Org. Advocacy | 72 | Researcher/Academic | 37 |
| Physician | 15 | Doctor | 70 | Physician | 37 |
| Researcher/Academic | 12 | Doctor | 60 | Doctor | 36 |
| Org. Advocacy | 10 | Patient Advocate | 56 | Professional Org. | 33 |
| Professional Org. | 8 | Individual Other Health | 55 | Researcher/Academic | 28 |
| Professional Org. | 7 | HCP | 49 | Researcher/Academic | 28 |
| Physician | 7 | Org. Advocacy | 42 | Physician | 27 |
| Individual Other Health | 7 | Researcher/Academic | 38 | Org. Advocacy | 25 |
